# Generation time of the Alpha and Delta SARS-CoV-2 variants

**DOI:** 10.1101/2021.10.21.21265216

**Authors:** WS Hart, E Miller, NJ Andrews, P Waight, PK Maini, S Funk, RN Thompson

## Abstract

**Background:** In May 2021, the Delta SARS-CoV-2 variant became dominant in the UK. This variant is associated with increased transmissibility compared to the Alpha variant that was previously dominant. To understand ongoing transmission and interventions, a key question is whether the Delta variant generation time (the time between infections in infector-infectee pairs) is typically shorter–i.e., transmissions are happening more quickly–or whether infected individuals simply generate more infections.

**Methods:** We analysed transmission data from a UK Health Security Agency household study. By fitting a mathematical transmission model to the data, we estimated the generation times for the Alpha and Delta variants.

**Results:** The mean intrinsic generation time (the generation time if there had been a constant supply of susceptibles throughout infection) was shorter for the Delta variant (4·6 days, 95% CrI 4·0-5·4 days) than the Alpha variant (5·5 days, 95% CrI 4·6-6·4 days), although within uncertainty ranges. However, there was a larger difference in the realised mean household generation time between the Delta (3·2 days, 95% CrI 2·4-4·2 days) and Alpha (4·5 days, 95% CrI 3·7-5·4 days) variants. This is because higher transmissibility led to faster susceptible depletion in households, in addition to the reduced intrinsic generation time.

**Conclusions:** The Delta variant transmits more quickly than previously circulating variants. This has implications for interventions such as contact tracing, testing and isolation, which are less effective if the virus is transmitted quickly. Epidemiological models of interventions should be updated to include the shorter generation time of the Delta variant.

## Introduction

In mid-2021, the Delta SARS-CoV-2 variant became the dominant variant in the UK^1^ and globally.^2^ This variant presents a higher risk of severe disease compared to previously circulating variants,^2,3^ although vaccination still provides significant protection.^4^ Concerningly, the Delta variant led to an increase in the growth rate of COVID-19 cases in the UK, swiftly outcompeting other SARS-CoV-2 variants.^1,5^ This was attributed to the increased transmissibility of the Delta variant, with an epidemiological study^5^ suggesting that it is 43-68% more transmissible than the Alpha variant that was identified in the UK in late 2020.^6^

Transmission of a SARS-CoV-2 variant can be characterised in two complementary ways: speed and strength.^7,8^ Speed refers to how quickly the variant grows at the population level, which is measured by the exponential growth rate and can be inferred from disease incidence data for that variant.^8^ However, understanding transmission of a novel variant also requires its strength to be estimated^8^ – this is the variant’s transmissibility, which is typically measured by the time-dependent reproduction number, or “R number” (the number of people that each infected person is expected to infect). The generation time (the time between infection events in infector-infectee pairs) determines the relationship between the speed and strength of a variant,^8–11^ and is a crucial input to many models used to estimate the reproduction number from case notification data.^12,13^ In principle, an increase in the growth rate of COVID-19 cases as observed for the Delta variant can be attributed to either an increase in transmissibility or a shortening of the generation time, or both of these factors.^8,14^

Previous studies have estimated the generation time for SARS-CoV-2 (for example ^15–18^), with most of these estimates using data collected during the very early months of the COVID-19 pandemic. A household study conducted by Public Health England (PHE) between February and November 2020 in the UK indicated that the generation time became shorter in the autumn of 2020 compared to earlier months.^18^ Since the SARS-CoV-2 generation time is changing, up-to-date generation time estimates are crucial to inform estimates of the reproduction number and to understand the relative transmissibility of different variants. However, the Alpha variant was responsible for infections in only two households in that study, while the Delta variant had not yet emerged by the end of the study.^18^ The effect of different variants on the SARS-CoV-2 generation time has therefore not been properly assessed.

Here, we report analyses of data from an ongoing household study conducted by the UK Health Security Agency (UKHSA; formerly PHE). As the study began in February 2021, there was an opportunity to estimate the generation time for both the Alpha and Delta variants and to analyse the transition from the Alpha variant to the Delta variant being dominant. We estimated two quantities for each variant. First, we estimated the *intrinsic generation time* (the generation time if the supply of susceptible individuals remains constant throughout infection). This quantity is widely applicable, since its distribution describes the relative transmission risk posed by an infected host at each time since infection, independently of the household structure. Second, we estimated the *household generation time* (the realised generation time in the study households). The mean household generation time is always shorter than the mean intrinsic generation time. This is because the household generation time reflects the fact that, in small household populations, infected individuals run out of susceptible individuals to infect.^9,19^ Most realised transmissions therefore happen more quickly within households than the intrinsic generation time distribution suggests.

In addition to estimating the household generation time for the Alpha and Delta variants, we estimated this quantity for individuals of different ages and vaccination statuses, and at different times during the study period. We found that the variant responsible for the infections within households had a larger effect on the household generation time than any of these other factors. This indicates that the increased growth rate of COVID-19 cases due to the Delta variant can be attributed to both an increase in transmissibility and a shortening of the generation time.

## Methods

### Data

We analysed household transmission data from the UKHSA HOCO2 study (Supplementary Data). This study began in February 2021 and is ongoing, and data were available from 227 households consisting of 557 participants at the time of our analyses (September 2021). Households were recruited to the study after an index case returned a positive PCR test, and (in most cases) three further PCR tests were subsequently carried out on each participating household member to determine whether they became infected. The following data were also collected from each participating household member:

- The date of any previous positive test result, where available.
- Symptom onset dates (for individuals who developed symptoms).
- The number of vaccine doses received, the vaccine type, and the date(s) of vaccination.

We assumed individuals were infected during the household transmission cluster if they returned a positive PCR test (including tests taken up to 28 days before the household was recruited to the study) and/or developed symptoms. Otherwise, we assumed individuals remained uninfected if they returned only negative tests. Household members who did not participate in the study, or withdrew before either taking a test or developing symptoms, were excluded from our analyses (but were included in the household size).

Genomic sequencing was carried out in 60% of study households. Where sequencing data were not available, we assumed that the Alpha variant was responsible for infections in households that were recruited to the study before May 2021, and the Delta variant for infections in households recruited after May 2021 (data from three households recruited during May 2021 in which sequencing was not carried out were excluded from our analyses).^1^ Overall, the Alpha and Delta variants were determined to be responsible for infections in 131 and 96 households, respectively.

### Mathematical modelling

We provide an overview of the model and the parameter inference approach here. Full details are given in the Supplementary Material.

We estimated the generation time from the household transmission data using a mechanistic approach motivated by compartmental modelling.^17,18^ In the model, each infection is divided into three successive, independently distributed, stages: latent, presymptomatic infectious and symptomatic infectious. Unlike commonly used approaches for estimating the SARS-CoV-2 generation time,^15,16^ this approach explicitly links the individual infectiousness profile of an infected host to exactly when they develop symptoms.

Data augmentation MCMC was used to fit the transmission model to the household data.^18,20^ We made the following assumptions when fitting the model to data:

- An incubation period distribution with mean 5.8 days and standard deviation 3.1 days.^21^
- Entirely asymptomatic infected hosts are 35% as infectious as those who develop symptoms.^22^
- Reduced susceptibility of vaccinated individuals compared to unvaccinated individuals (a ‘leaky’ vaccine was assumed), using estimates of protection against infection obtained by Pouwels and colleagues (Table S1).^23^
- All household transmission clusters in the study originated with a single primary case, with no further infections introduced into the household from outside.

Values of four model parameters for each of the Alpha and Delta variants were estimated in the parameter fitting procedure:

- The mean latent period (the time from being infected to becoming infectious), as a proportion of the mean incubation period.
- The mean symptomatic infectious period.
- The relative infectiousness of presymptomatic infectious hosts compared to those with symptoms.
- The overall transmissibility parameter, *β*_0_, which describes the expected number of household transmissions generated by a single infector (who develops symptoms) in a large, otherwise entirely susceptible and unvaccinated, household.

Posterior distributions of fitted model parameters are shown in Figure S1.

The intrinsic generation time distribution was calculated for each variant from the assumed and estimated values of model parameters. Realised generation times within the study households were also estimated at each step of the MCMC procedure. This enabled us to compare household generation times based on variant, vaccination status, date and age.

## Results

In Figure 1A, we show posterior estimates of the mean intrinsic generation time (which involves an assumption that infected individuals have contacts with susceptible hosts at a constant rate throughout their infectious period, and therefore provides a generalised generation time estimate that is independent of the household size) for the Alpha and Delta variants. We estimated this quantity to be slightly shorter for the Delta variant (4·6 days, 95% CrI 4·0-5·4 days) compared to the Alpha variant (5·5 days, 95% CrI 4·6-6·4 days), although there was overlap between the credible intervals (Figure 1A). On the other hand, our results indicate a substantially higher transmissibility within households of the Delta variant compared to the Alpha variant (Figure 1B).

**Figure 1.**
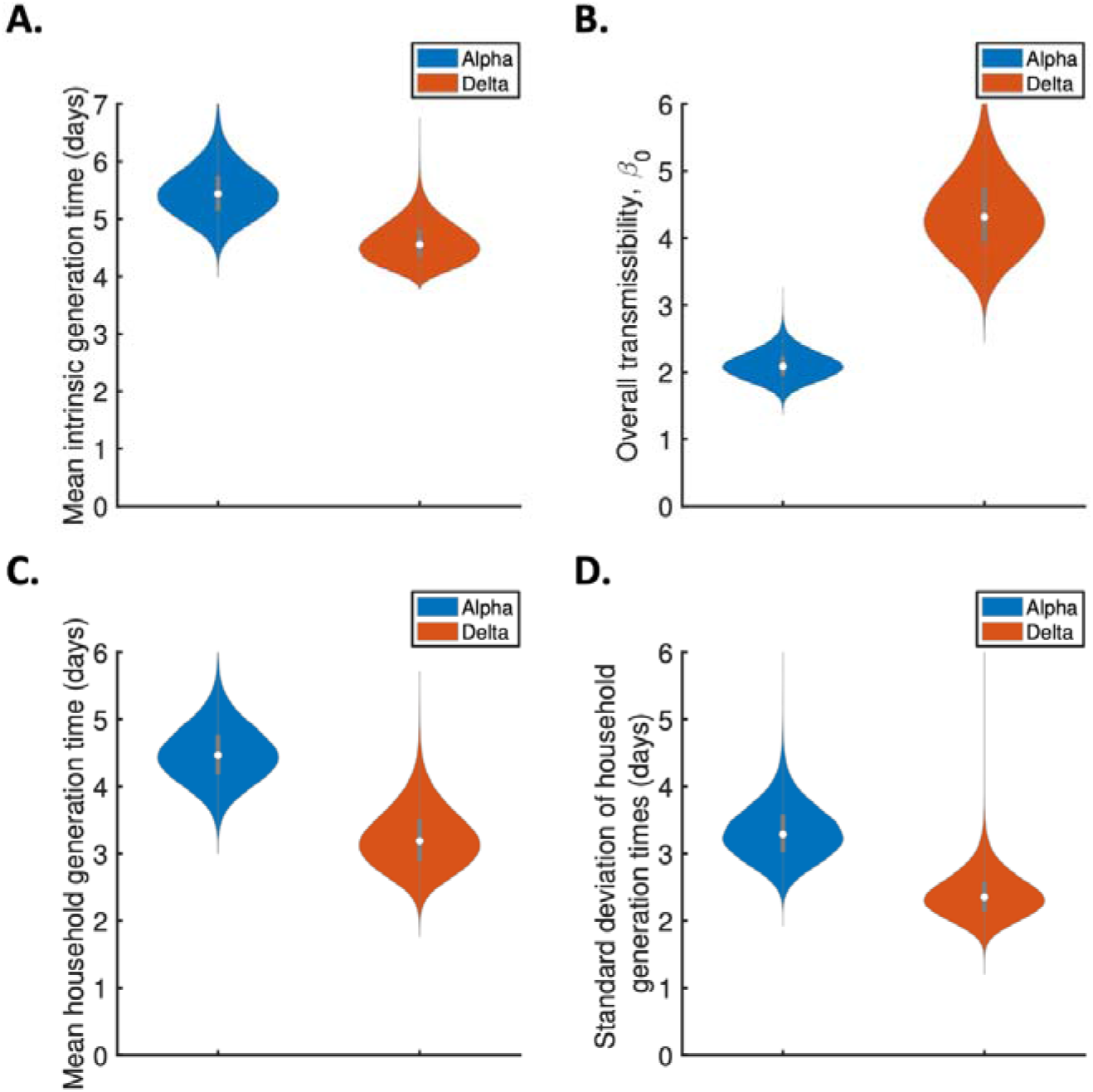
The effect of variant on the intrinsic generation time and the household generation time. Violin plots indicate posterior estimates for the Alpha (blue) and Delta (red) variants of: A. The mean intrinsic generation time (i.e., the mean generation time if the supply of susceptibles remains constant throughout infection); B. The overall transmissibility parameter, *β*_0_ (i.e., the expected number of household transmissions generated by a single infected individual introduced into a large, entirely susceptible, household); C. The mean household generation time (i.e., the mean generation time accounting for susceptible depletion in the households); D. The standard deviation of household generation times. Posterior means and 95% credible intervals for these quantities are given in Table S2 and Table S3 in the Supplementary Material.

Estimates of the mean household generation time for the two variants (Figure 1C) are shorter than the corresponding intrinsic generation time estimates (Figure 1A), since the household generation time accounts for susceptible depletion within households (which means that potential transmissions with longer generation times are less likely to occur). We found a shorter mean household generation time for the Delta variant (3·2 days, 95% CrI 2·4-4·2 days) than for the Alpha variant (4·5 days, 95% CrI 3·7-5·4 days). This is due to the higher transmissibility of the Delta variant (Figure 1B) leading to faster susceptible depletion, in addition to the smaller reduction in the intrinsic generation time shown in Figure 1A. We also estimated the standard deviation of generation times within households to be smaller for the Delta variant than for the Alpha variant (Figure 1D). In Figure S2 of the Supplementary Material, we combined the estimates obtained in each step of the data augmentation MCMC parameter fitting procedure (which are summarised in Figure 1) to obtain an estimate of the entire household generation time distribution. This also indicates that household transmissions with the Delta variant typically occur earlier in infection (Figure S2C).

We note that the estimates in Figure 1B cannot be compared directly with previous estimates of the transmission advantage of the Delta variant (e.g., an estimate of 43-68% obtained using incidence data^5^), since the value of *β*_0_ reflects the instrinsic transmissibility of the Alpha and Delta variants rather than realised transmission. The intrinsic transmission advantage of the Delta variant will only be realised if sufficiently many susceptibles are available to each infector.

We also explored the effects of vaccination (Figure 2A-C), age (Figure 2D-E) and month of recruitment to the household study (Figure 2F) on household generation times. We estimated the generation time associated with infectors (Figure 2A) and infectees (Figure 2B) of different vaccination status, as well as considering infector-infectee pairs with different combinations of vaccine status (Figure 2C), each time stratifying our estimates by variant. In each case, the effect of variant on the household generation time was substantially larger than the effect of vaccination status (Figure 2A-C). Similarly, for both variants there was no clear effect of the age of infectors (Figure 2D) or infectees (Figure 2E) on the generation time. Finally, a reduction in the household generation time occurred between April and June 2021 (Figure 2F), likely due to the Delta variant becoming dominant in the UK during May 2021.^1^

**Figure 2.**
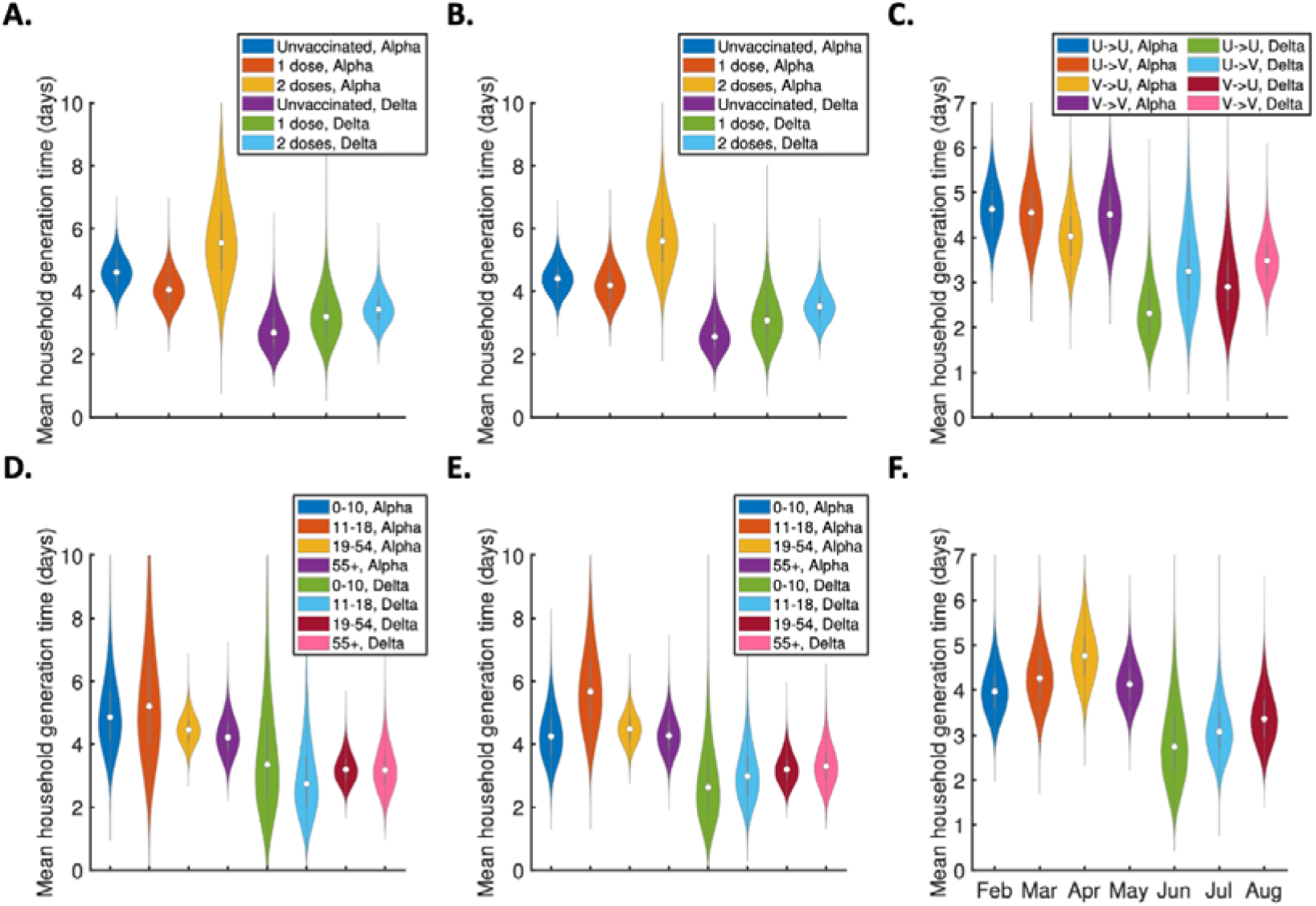
The effect of different factors on household generation times. Violin plots indicate posterior estimates of the mean household generation time for infectors, infectees and households with different characteristics, showing comparisons based on: A. Vaccination status of the infector and variant; B. Vaccination status of the infectee and variant; C. Combination of vaccination statuses in the infector-infectee pair (for example, U->V corresponds to transmissions from unvaccinated infectors to vaccinated infectees) and variant; D. Age of the infector and variant; E. Age of the infectee and variant; F. Recruitment month of the household into the study (all months shown are in 2021).

## Discussion

Novel SARS-CoV-2 variants possess different transmission characteristics to the virus that originally emerged in Wuhan, China.^5,14^ A key transmission characteristic is the generation time, which measures the speed of transmission between individuals.^17,19,24^ To estimate the generation time, it is necessary to analyse datasets in which it is possible to determine or estimate who-infected-whom.^17,19,24^ Household studies provide an opportunity to conduct such analyses.^18,20^ Here, we have analysed data from the HOCO2 household study conducted by the UKHSA, and compared the generation time for the Alpha and Delta variants (which were the dominant variants in England at the beginning and end of the time period considered^1^).

We first estimated the intrinsic generation time, which describes the generation time under the assumption that there is a steady supply of susceptible individuals available to infect (i.e., susceptible depletion in households is not accounted for). The mean intrinsic generation time was found to be slightly lower for the Delta variant than for the Alpha variant (Figure 1A), but there was a substantial overlap in the credible intervals between the two variants. However, the data indicated that the Delta variant is much more transmissible in households than the Alpha variant (Figure 1B). The impact of this increased transmissibility is a larger difference in realised generation times within households between the two variants (Figure 1C) than would be expected from the difference in the intrinsic generation time alone. This is because increased transmissibility leads to susceptible individuals in the household being infected quickly, thereby being unavailable for infection later (thus shortening the average period between realised transmissions). The effect of variant on the household generation time was more substantial than the effects of other factors, including age and vaccination status (Figure 2).

Our study is the first to compare the generation time for different SARS-CoV-2 variants directly, and we are not aware of any other generation time estimate for the Alpha variant. A generation time estimate for the Delta variant based on 55 transmission pairs was recently stated in China CDC Weekly,^25^ although the methods underlying that estimate were not presented. Nonetheless, the mean generation time estimate of 2·9 days in that study^25^ lies within the credible interval of our estimate of the mean household generation time for the Delta variant (3·2 days, 95% CrI 2·4-4·2 days) and is shorter than the lower credible limit of our estimate for the Alpha variant (4·5 days, 95% CrI 3·7-5·4 days). This supports our finding that the Delta variant is transmitted more quickly than the Alpha variant.

Other studies have estimated quantities related to the generation time for the Alpha and Delta variants, including the serial interval^26–28^ and the viral load trajectories of infected hosts.^29–31^ However, while the serial interval (the interval between symptom onset times in infector-infectee pairs) is sometimes used as a proxy for the generation time (the interval between infection times), these two quantities may not follow the same distribution^24^ – for example, the generation time always takes positive values, whereas the serial interval can be negative for pathogens such as SARS-CoV-2 for which presymptomatic transmission can occur – and may not even have the same mean value.^18^ Similarly, although it may be possible to infer how infectiousness varies during infection based on viral load data,^32^ the timing of realised transmissions also depends on other factors as well as intrinsic infectiousness, such as behaviour (for example, individuals may be less likely to transmit after developing symptoms and isolating) and the availability of susceptibles. Our approach explicitly accounts for changes in the transmission risk following symptom onset, while our estimates of the household generation time account for depletion of susceptibles within households.

Estimates of the generation time underlie a range of epidemiological analyses, including estimation of the time-varying reproduction number (or “R number”)^12,13^ in different regions or countries and the relative transmissibility of different variants^5,14^. Such analyses often neglect temporal changes in the generation time, but our results indicate that the generation time is shorter for the Delta variant than for the Alpha variant. When estimating reproduction numbers, overestimation of the mean generation time generally leads to reproduction number estimates that are further from one (the threshold for an outbreak being under control) compared to the true value.^10^ When analysing the transmissibility advantage of the Delta variant over the Alpha variant, neglecting a shorter generation time for the Delta variant would lead to its transmissibility advantage being overestimated.^8^ We contend that the observed growth rate advantage of the Delta variant is likely due to a combination of both increased transmissibility and a shorter generation time.

Since susceptible depletion during infection may be a less important determinant of the timing of transmissions occurring outside households than within households, we expect the overall generation time distribution (accounting for transmissions across all settings) to lie somewhere between our “household” and “intrinsic” estimates. This suggests that inferring the transmissibility advantage of the Delta variant over the Alpha variant from incidence data using our household generation time estimates (which show a greater difference between variants) would give a lower bound for the transmissibility advantage (since the observed increased growth rate of the Delta variant is then explained in part by a substantial generation time reduction). In contrast, we expect that inferring the transmissibility advantage of the Delta variant over the Alpha variant using the intrinsic generation time estimates would give an upper bound.

In summary, our analyses suggests that the Delta variant transmits more quickly than the previously circulating Alpha variant. This has implications for interventions, since public health measures such as contact tracing are less effective if transmission occurs quickly. It is essential for public health that epidemiological models are updated to reflect the generation time of the variants driving transmission, and that the generation time is assessed in future as the characteristics of SARS-CoV-2 transmission continue to change.

## Data Availability

All data produced are available online at https://github.com/will-s-hart/variant_generation_times.

## Data sharing

All data generated or analysed during this study are available in this published article and its supplementary files. Data and code for reproducing our results are available at https://github.com/will-s-hart/variant_generation_times.

## Acknowledgements

The data collection for the study was undertaken by the UKHSA (an executive agency of the Department of Health) as part of the COVID-19 response. WSH was funded by an EPSRC Excellence Award for his doctoral studies (Grant Reference EP/R513295/1). EM receives support from the National Institute for Health Research (NIHR) Health Protection Research Unit in Immunisation at the London School of Hygiene and Tropical Medicine in partnership with the UKHSA (Grant Reference NIHR200929). SF receives funding from the Wellcome Trust (Grant Reference 210758/Z/18/Z). RNT receives support from the UKRI (Grant Reference EP/V053507/1). Thanks to the household members who took part in this study, the nursing staff at the UKHSA who recruited and followed up the households, the laboratory staff at the UKHSA who tested the swabs and the UKHSA administrative staff who arranged for the delivery and collection of testing kits from households. The household surveillance protocol was approved by the UKHSA Research Ethics and Governance Group as part of the portfolio of the UKHSA’s enhanced surveillance activities in response to the pandemic. Oral informed consent for sampling and follow up was obtained by the nurses from household members who were free to decline to participate in the surveillance at any time. Consent for children was obtained by a parent or legal guardian. Only anonymised data were provided to non-UKHSA authors.

## Authors’ contributions

Conceptualization: WSH, SF, RNT; Literature search: WSH, RNT; Data collection and curation: EM, NJA, PW; Formal analysis: WSH; Investigation: WSH; Methodology: WSH, SF, RNT; Project administration: SF, RNT; Software: WSH; Supervision: PKM, SF, RNT; Validation: WSH; Visualization: WSH; Writing – original draft: WSH, RNT; Writing – review & editing: all authors.

