## Supplementary Material for "Generation time of the Alpha and Delta SARS-CoV-2 variants"

### Supplementary Methods

#### General modelling framework

We assumed the expected force of infection, $\beta(\tau)$, exerted by an infected host onto each susceptible member of their household at time 𝜏 since infection to be given by

$$\beta(\tau)=\frac{\beta_{0}}{(n-1)}f(\tau),$$

for a host who develops symptoms, and

$$\beta(\tau)=\frac{\alpha_{A}\beta_{0}}{(n-1)}f(\tau),$$

for a host who remains asymptomatic throughout infection. Here:

- $\beta_{0}$ represents the overall transmissibility for a host who develops symptoms.
- $n$ is the household size (we assumed frequency-dependent household transmission).
- $f(\tau)$ is the intrinsic generation time distribution (i.e., the generation time distribution in the absence of susceptible depletion during the course of infection).
- $\alpha_{A}$ is the relative infectiousness of infected hosts who remain asymptomatic throughout infection (compared to infected hosts who develop symptoms). We assumed a value of $\alpha_{A}=0\text{⋅}35$.^1^

The (expected) instantaneous probability density of the infected host under consideration infecting a given susceptible household member (denoted $j$) is given by $\eta_{j}\beta(\tau)$, where $\eta_{j}$ describes the relative susceptibility of individual $j$. This susceptibility was assumed to depend on vaccination status according to previous estimates of vaccine efficacy against infection by the Alpha and Delta variants (see Table S1).^2^

In our approach for estimating the generation time, infectiousness is explicitly linked to symptoms, so that the infectiousness profile of a given infector depends on exactly when they develop symptoms. Throughout, the expected force of infection exerted by an infected host onto each susceptible member of their household at time 𝜏 since infection, conditional on incubation period $\tau_{inc}$, is denoted by $\beta(\tau\mid\tau_{inc})$.

#### Individual infectiousness model

We considered a mathematical model in which each infected host (who develops symptoms) progresses through independent latent (*E*), presymptomatic infectious (*P*) and symptomatic infectious (*I*) stages of infection. The transmission rates of the host during the *P* and *I* stages are denoted by $\beta_{P}$ and $\beta_{I}$, respectively, and we denote their ratio $\alpha_{P}=\beta_{P}/\beta_{I}$. We assumed the duration of each stage, denoted $y_{E/P/I}$, to be gamma distributed:

$$y_{E}\sim\mathrm{Gamma}(k_{E},1/{(k}_{inc}\gamma)),$$

$$y_{P}\sim\mathrm{Gamma}(k_{P},1/{(k}_{inc}\gamma)),$$

$$y_{I}\sim\mathrm{Gamma}(k_{I},1/{(k}_{I}\mu)),$$

where we write $X\sim Gamma(a,b)$ for a gamma distributed random variable with shape parameter $a$ and scale parameter $b$. We assumed that $k_{E}+k_{P}=k_{inc}$, so that the incubation period, $\tau_{inc}=y_{E}+y_{P}$, is gamma distributed, with

$$\tau_{inc}\sim\mathrm{Gamma}\left( k_{inc},1/{(k}_{inc}\gamma) \right).$$

Hosts who remain asymptomatic throughout infection were assumed to follow the same *E*/*P*/*I* stages, although in this case the distinction between the *P* and *I* stages has no epidemiological meaning. Stage durations, as well as the value of $\alpha_{P}$, were assumed to be identical for entirely asymptomatic hosts and those who develop symptoms. Similarly, vaccination was not assumed to affect model parameters (other than the relative susceptibility of vaccinated hosts), although we explored the effect of vaccination on realised household generation times in Figure 2.

We fixed the values of the parameters $k_{inc}$ and $1/\gamma$ (which represent the shape parameter of the incubation period distribution and the reciprocal of the mean incubation period, respectively) in order to obtain an incubation period distribution of mean 5·8 days and standard deviation 3·1 days.^3^ The values of $k_{inc}$ and $1/\gamma$ are given in Table S1. We assumed that $k_{I}=1$, so the symptomatic infectious period is exponentially distributed. The following quantities were then estimated for the Alpha and Delta variants when we fitted the model to the household transmission data:

- The ratio between the mean latent (*E*) period and the mean incubation (combined *E* and *P*) period, $k_{E}/k_{inc}$.
- The mean symptomatic infectious (*I*) period, $1/\mu$.
- The ratio between the transmission rates when potential infectors are in the *P* and *I* stages, $\alpha_{P}$.
- The overall transmissibility parameter, $\beta_{0}$.

*Conditional infectiousness*

For a host who develops symptoms, conditional on incubation period $\tau_{inc}$, their expected infectiousness at time since infection $\tau$ is^4^

$$\beta(\tau\mid\tau_{inc})=\left\{ \begin{matrix} \frac{\alpha_{P}C\beta_{0}}{(n-1)}\left( 1-F_{Beta}(1-\tau/\tau_{inc};k_{P},k_{E}) \right), & 0<\tau<\tau_{inc}, \\ \frac{C\beta_{0}}{(n-1)}\left( 1-F_{I}(\tau-\tau_{inc}) \right), & \tau>\tau_{inc}. \end{matrix} \right.$$

Here, $\beta_{0}$ is the overall transmissibility parameter, $n$ is the household size, $F_{I}(y)$ is the cumulative distribution of the duration of the *I* stage, $F_{Beta}(x;a,b)$ is the cumulative distribution of a beta distributed random variable with shape parameters $a$ and $b$, and

$$C=\frac{k_{inc}\gamma\mu}{\alpha_{P}k_{P}\mu+k_{inc}\gamma}.$$

The cumulative conditional infectiousness up to time $\tau$ can therefore be calculated to be

$$B\left( \tau\mid\tau_{inc} \right)=\int_{0}^{\tau} \beta\left( \tilde{\tau} \mid\tau_{inc} \right)d\tilde{\tau} =\left\{ \begin{aligned} \left( \tau-\tau_{inc} \right)\beta\left( \tau\mid\tau_{inc} \right)+\frac{\alpha_{P}C\beta_{0}}{(n-1)}\left[ \frac{k_{p}\tau_{inc}}{k_{inc}}\left( 1-F_{Beta}\left( 1-\tau/\tau_{inc};k_{P}+1,k_{E} \right) \right) \right], &0\leq\tau<\tau_{inc}, \\ \left( \tau-\tau_{inc} \right)\beta\left( \tau\mid\tau_{inc} \right)+\frac{C\beta_{0}}{(n-1)}\left[ \frac{{\alpha k}_{p}\tau_{inc}}{k_{inc}}+\frac{1}{\mu}F_{Gamma}\left( \tau-\tau_{inc};k_{I}+1,\frac{1}{k_{I}\mu} \right) \right], &\tau\geq\tau_{inc}, \end{aligned} \right.$$

where $F_{Gamma}(x;a,b)$ is the cumulative distribution of a gamma distributed random variable with shape parameter $a$ and scale parameter $b$. The total force of infection on each household member (over the infector’s course of infection) is then

$$B\left( \infty\mid\tau_{inc} \right)=\frac{\beta_{0}}{(n-1)}\left( \frac{\alpha_{P}k_{P}\gamma\mu\tau_{inc}+k_{inc}\gamma}{\alpha_{P}k_{P}\mu+k_{inc}\gamma} \right).$$

The mean of this expression over the incubation period distribution is $\frac{\beta_{0}}{(n-1)}$.

For a host who remains asymptomatic throughout infection, conditional on the combined duration of the *E* and *P* stages, $\tau_{inc}=y_{E}+y_{P}$, the infectiousness, $\beta\left( \tau\mid\tau_{inc} \right)$, is given by the product of $\alpha_{A}$ and the corresponding expression for a host who develops symptoms. We note that in this case, $\tau_{inc}$ has no epidemiological interpretation, but this formulation was convenient when fitting the model to data (see “Parameter fitting” below).

*Intrinsic generation time distribution*

The generation time, $\tau_{gen}$, can be written as

$$\tau_{gen}=y_{E}+y^{*},$$

where $y_{E}$ is the length of the latent (*E*) stage, and $y^{*}$ is the time from the start of the presymptomatic infectious (*P*) stage to the transmission occurring. As shown in our previous work,^4^ if the effect of susceptible depletion during infection is neglected, $y^{*}$ has density

$$f^{*}\left( y^{*} \right)=C\left( \alpha_{P}\left( 1-F_{P}\left( y^{*} \right) \right)+\int_{0}^{y^{*}} \left( 1-F_{I}\left( y^{*}-y_{P} \right) \right)f_{P}\left( y_{P} \right)dy_{P} \right).$$

Using this density, it can be shown that the moments of this distribution are

$$E\left[ \left( y^{*} \right)^{m} \right]=\frac{C}{m+1}\left( \alpha_{P}E\left[ {y_{P}}^{m+1} \right]+E\left[ \left( y_{P}+y_{I} \right)^{m+1}{{-y}_{P}}^{m+1} \right] \right).$$

In particular,

$$E\left[ y^{*} \right]=\frac{C}{2}\left( \alpha_{P}E\left[ {y_{P}}^{2} \right]+2E\left[ y_{P} \right]E\left[ y_{I} \right]+E\left[ {y_{I}}^{2} \right] \right),$$

and

$$\mathrm{Var}\left[ y^{*} \right]=\frac{C}{3}\left( \alpha_{P}E\left[ {y_{P}}^{3} \right]+3E\left[ {y_{P}}^{2} \right]E\left[ y_{I} \right]+3E\left[ y_{P} \right]E\left[ {y_{I}}^{2} \right]+E\left[ {y_{I}}^{3} \right] \right)-\left( E\left[ y^{*} \right] \right)^{2}.$$

Note that for a gamma distributed random variable, $X\sim\mathrm{Gamma}(a,b)$, we have

$$E\left[ X^{m} \right]=\frac{\Gamma\left( a+m \right)}{\Gamma\left( a \right)}b^{m}=a\left( a+1 \right)\ldots\left( a+\left( m-1 \right) \right)b^{m}.$$

Therefore, for gamma distributed stage durations, explicit expressions can be obtained for the mean and variance of the intrinsic generation time distribution,

$$E\left[ \tau_{gen} \right]=E\left[ y_{E} \right]+E\left[ y^{*} \right],$$

$$\mathrm{Var}\left[ \tau_{gen} \right]=\mathrm{Var}\left[ y_{E} \right]+\mathrm{Var}\left[ y^{*} \right],$$

where the latter expression holds because $y_{E}$ and $y^{*}$ are assumed to be independent.

#### Likelihood function

Here, we consider a household of size $n$, in which $n_{I}$ household members become infected (of whom $n_{S}$ develop symptoms and $n_{A}$ remain asymptomatic throughout infection) and $n_{U}=n-n_{I}$ remain uninfected. We derive an expression for the likelihood of the vector of unknown model parameters,

$$\theta=\left( k_{E}/k_{inc},1/\mu,\alpha_{P},\beta_{0} \right),$$

(where $\theta$ was assumed to be different for the Alpha and Delta variants), given:

i. The entire sequence of infection times of individuals in the household ($t_{1}<\ldots<t_{n_{I}}$).
ii. The precise symptom onset time ($t_{s,j}$) of each host, $j$, who develops symptoms.
iii. The times at which entirely asymptomatic infected hosts enter the *I* stage of infection (also denoted by $t_{s,j}$).

Since exact infection and symptom onset times were not available within study households, we used data augmentation MCMC to fit the two models to the household transmission data using this likelihood function (see further details below).

When deriving the likelihood, we made the following simplifying assumptions:

- The virus is introduced once into the household (i.e., no subsequent infections from the community occur following the infection of the primary case).
- No co-primary cases.
- Potential bias towards more recent infection of the primary host if community prevalence is increasing, or less recent if prevalence is decreasing,^5–7^ was neglected.

We denote the conditional infectiousness of household member $j$, at time $\tau$ since infection, by $\beta_{j}(\tau\mid t_{s,j}-t_{j})$, where ${(t}_{s,j}-t_{j}$) corresponds to the incubation period for a host who develops symptoms. The total (instantaneous) force of infection exerted at time $t$ on each susceptible household member is then

$$\lambda(t)=\sum_{j=1}^{n_{I}} \beta_{j}(t-t_{j}\mid t_{s,j}-t_{j}),$$

where $\beta_{j}\left( t-t_{j}\mid t_{s,j}-t_{j} \right)=0$ for $t\leq t_{j}$, and the cumulative force of infection is

$$\Lambda\left( t \right)=\int_{-\infty}^{t} \lambda\left( s \right)ds=\sum_{j=1}^{n_{I}} B_{j}(t-t_{j}\mid t_{s,j}-t_{j}),$$

where $B_{j}(\tau\mid t_{s,j}-t_{j})$ denotes the cumulative conditional infectiousness of individual $j$.

We assumed that the probability of each host being the one who introduced the virus into the household to be proportional to their relative susceptibility, leading to a likelihood contribution of

$$\frac{\eta_{1}}{\sum_{j=1}^{n} \eta_{j}},$$

where $\eta_{j}$ denotes the relative susceptibility of host $j$. For $k=2,\ldots,n_{I}$, conditional on the sequence of infection times up to time $t_{k}$, the probability of host $k$ becoming infected at time $t_{k}$ is given by

$$\eta_{k}\lambda\left( t_{k} \right)\exp\left( -\eta_{k}\Lambda\left( t_{k} \right) \right),$$

where $\exp(-\eta_{k}\Lambda(t_{k}))$ represents the probability of host $k$ avoiding infection up to time $t$.^8,9^ For $k=n_{I}+1,\ldots,n$, conditional on the entire sequence of infection times, $t_{1},\ldots{,t}_{n_{I}}$, the probability of host $k$ remaining uninfected is given by $\exp(-\eta_{k}\Lambda(\infty))$.

The likelihood, $L\left( \theta\right)$, can therefore be written as

$$L\left( \theta\right)=\prod_{k=1}^{n} L_{k,1}\left( \theta\right)L_{k,2}\left( \theta\right).$$

Here, $L_{k,1}(\theta)$ is the contribution to the likelihood from the transmission, or absence of transmission, to host $k$, i.e.,

$$L_{k,1}(\theta)=\left\{ \begin{aligned} \frac{\eta_{1}}{\sum_{j=1}^{n} \eta_{j}}, &\mathrm{for}k=1; \\ \eta_{k}\lambda(t_{k})exp(-\eta_{k}\Lambda(t_{k})), &\mathrm{for}k=2,\ldots,n_{I}; \\ \exp(-\eta_{k}\Lambda(\infty)), &\mathrm{for}k=n_{I}+1,\ldots,n. \end{aligned} \right.$$

For an infected host $k$, $L_{k,2}(\theta)$ is the likelihood contribution from their incubation period (or for an entirely asymptomatic infected host, the corresponding combined duration of the *E* and *P* stages of infection), i.e.,

$$L_{k,2}(\theta)=\left\{ \begin{aligned} f_{inc}(t_{s,k}-t_{k}), &\mathrm{for}k=1,\ldots,n_{I}; \\ 1, &\mathrm{for}k=n_{I}+1,\ldots,n. \end{aligned} \right.$$

where $f_{inc}$ is the probability density function of the incubation period.

#### Parameter estimation

Unknown model parameters were estimated for each variant using data augmentation MCMC. The observed transmission data comprised information about whether or not individuals were ever infected and/or displayed symptoms, symptom onset dates, and for some individuals an upper bound on their infection time (corresponding to the date of a positive PCR test). These data were augmented with:

i. The infection time, $t_{j}$, of each infected host.
ii. The time, $t_{s,j}$, at which each infected host transitioned from the *P* stage to the *I* stage of infection (this corresponds to the symptom onset time for a host who developed symptoms).

No prior assumptions were made about the order of transmissions within the household.

For each variant, we assumed lognormal priors for $1/\mu$ (the mean duration of the symptomatic infectious period), $\alpha_{P}$ (the ratio of the transmission rates during the presymptomatic infectious and symptomatic infectious stages of infection) and $\beta_{0}$ (the overall transmissibility). Since $\alpha_{P}$ represents a ratio between transmission rates, a prior with median 0 was used to ensure equal prior probabilities of values above and below 1. This prior was also chosen to limit the prior probability of extreme values, with a prior 95% credible interval of [0·2,5]. A beta prior was used for $k_{E}/k_{inc}$ (the ratio of the mean durations of the latent and incubation periods, which was constrained to lie between 0 and 1), and was chosen to restrict the prior probability of values very close to either 0 or 1. All prior distributions were taken to be independent (between different parameters, and between variants). The exact priors we used are given in Table S2.

In the description of the parameter fitting procedure below, we denote the augmented data by

$$\boldsymbol{t}=\left( \boldsymbol{t}^{\left( 1 \right)},\ldots,\boldsymbol{t}^{\left( M \right)} \right),$$

where $\boldsymbol{t}^{\left( m \right)}$ represents the augmented data from household $m=1,\ldots,M$, and $M$ is the total number of households. We denote the vector of fitted parameters by $\theta$, where

$$\theta=(\theta^{(Alpha)},\theta^{(Delta)})$$

now includes the parameter values for both variants. We write the (overall) likelihood as

$$L\left( \theta;\boldsymbol{t} \right)=\prod_{m=1}^{M} L^{(m)}\left( \theta^{(V_{m})};\boldsymbol{t}^{(m\boldsymbol{)}} \right),$$

where $V_{m}$ denotes the variant responsible for infections in household $m$, and the likelihood contributions, $L^{(m)}\left( \theta^{(V_{m})};\boldsymbol{t}^{(m\boldsymbol{)}} \right)$, were computed as described in the previous section (i.e., all households in the study were assumed to be independent). Finally, we denote the prior density of $\theta$ by $\pi\left( \theta\right).$

In each step of the chain, we carried out (in turn) one of the following:

1. Propose new values for each entry of the vector of model parameters, $\theta$, using a multivariate normal proposal distribution (around the value of $\theta$ in the previous step of the chain; a correlation of 0·5 was used between the proposal distributions of $k_{E}/k_{inc}$ and $\alpha_{P}$, and between those of $1/\mu$ and $\alpha_{P}$, for each variant). Accept the proposed parameters, $\theta_{prop}$, with probability

$$\min\left( \frac{L\left( \theta_{prop};\boldsymbol{t} \right)\pi(\theta_{prop})}{L\left( \theta_{old};\boldsymbol{t} \right)\pi(\theta_{old})},1 \right),$$

where $\theta_{old}$ denotes the vector of parameter values from the previous step of the chain, and where the augmented data, $\boldsymbol{t}$**,** remain unchanged in this step.

2. Propose new values for the precise symptom onset times of each symptomatic infected host, using independent uniform proposal distributions (within the day of symptom of onset for each host). For each household, $m$, accept the proposed augmented data, $\boldsymbol{t}_{prop}^{(m\boldsymbol{)}}$, from that household with probability

$$\min\left( \frac{L^{(m)}\left( \theta^{(V_{m})};\boldsymbol{t}_{prop}^{(m\boldsymbol{)}} \right)}{L^{(m)}\left( \theta^{(V_{m})};\boldsymbol{t}_{old}^{(m\boldsymbol{)}} \right)},1 \right),$$

where $\boldsymbol{t}_{old}^{(m\boldsymbol{)}}$ denotes the corresponding augmented data from the previous step of the chain, and where the model parameters, $\theta$, remain unchanged in this step (i.e., proposed times are accepted/rejected independently for each household, according to the likelihood contribution from that household).

3. Propose new values for the infection time of one randomly chosen infected host in each household (either symptomatic or asymptomatic), using independent normal proposal distributions (around the equivalent times in the previous step of the chain). For each household, $m$, accept the proposed augmented data, $\boldsymbol{t}_{prop}^{(m\boldsymbol{)}}$, from that household with probability

$$\min\left( \frac{L^{(m)}\left( \theta^{(V_{m})};\boldsymbol{t}_{prop}^{(m\boldsymbol{)}} \right)}{L^{(m)}\left( \theta^{(V_{m})};\boldsymbol{t}_{old}^{(m\boldsymbol{)}} \right)},1 \right).$$

4. Propose new values for both the infection time, $t$, and the time of the start of the *I* stage, $t_{s}$, holding $t_{s}-t$ constant, for one randomly chosen asymptomatic infected host in each household (in households where there was at least one), using independent normal proposal distributions (around the equivalent times in the previous step of the chain). For each household, $m$, accept the proposed augmented data, $\boldsymbol{t}_{prop}^{(m\boldsymbol{)}}$, from that household with probability

$$\min\left( \frac{L^{(m)}\left( \theta^{(V_{m})};\boldsymbol{t}_{prop}^{(m\boldsymbol{)}} \right)}{L^{(m)}\left( \theta^{(V_{m})};\boldsymbol{t}_{old}^{(m\boldsymbol{)}} \right)},1 \right).$$

#### Sampling of household generation times

Realised household generation times may be shorter than predicted by the intrinsic generation time distribution, $f(\tau)$, due to the depletion of susceptible household members before longer generation times can be attained.^10–12^ For example, if infected hosts are (on average) equally infectious at two times since infection, $\tau_{1}<\tau_{2}$, then $f\left( \tau_{1} \right)=f\left( \tau_{2} \right)$. However, because the number of susceptible household members may decrease between these two times (i.e., either the host under consideration, or another infected household member, may transmit the virus within the household in the intervening time), then transmission is in fact more likely to occur in a household at the earlier time, $\tau_{1}$, when more susceptibles are available.

Therefore, we also estimated the realised generation times within the study households. These were sampled during the parameter fitting procedure as follows: in a given household, for $k>1$, the instantaneous probability of individual $k$ (the $k$^th^ household member to be infected) being infected at time $t_{k}$ is

$$\eta_{k}\lambda\left( t_{k} \right)=\eta_{k}\sum_{j=1}^{k-1} \beta_{j}(t_{k}-t_{j}\mid t_{s,j}-t_{j}),$$

where:

- $t_{1}<t_{2}<\ldots t_{(k-1)}$ are the infection times of the first $(k-1)$ individuals to be infected in the household.
- $t_{s,j}$ is the symptom onset time of individual $j$ (or the entry time into the *I* stage of infection for an entirely asymptomatic infected host).
- $\beta_{j}(\tau\mid t_{s,j}-t_{j})$ is the (conditional) infectiousness of individual $j$ at time since infection $\tau$.
- $\eta_{k}$ is the relative susceptibility of individual $k$.

Conditional on this transmission occurring at time $t_{k}$, the probability that host $j$ is responsible for the transmission (for $j=1,\ldots,(k-1)$) is given by

$$p_{j}=\frac{\beta_{j}\left( t_{k}-t_{j} \mid t_{s,j}-t_{j} \right)}{\lambda\left( t_{k} \right)},$$

i.e., the generation time corresponding to the $k$^th^ transmission is ${(t}_{k}-t_{j})$ with probability $p_{j}$.

During the parameter fitting procedure, we calculated the probabilities $p_{j}$ corresponding to each transmission given the augmented data. We used these probabilities to sample the infector responsible for each transmission and therefore obtain samples of estimated household generation times.

### Supplementary Figures

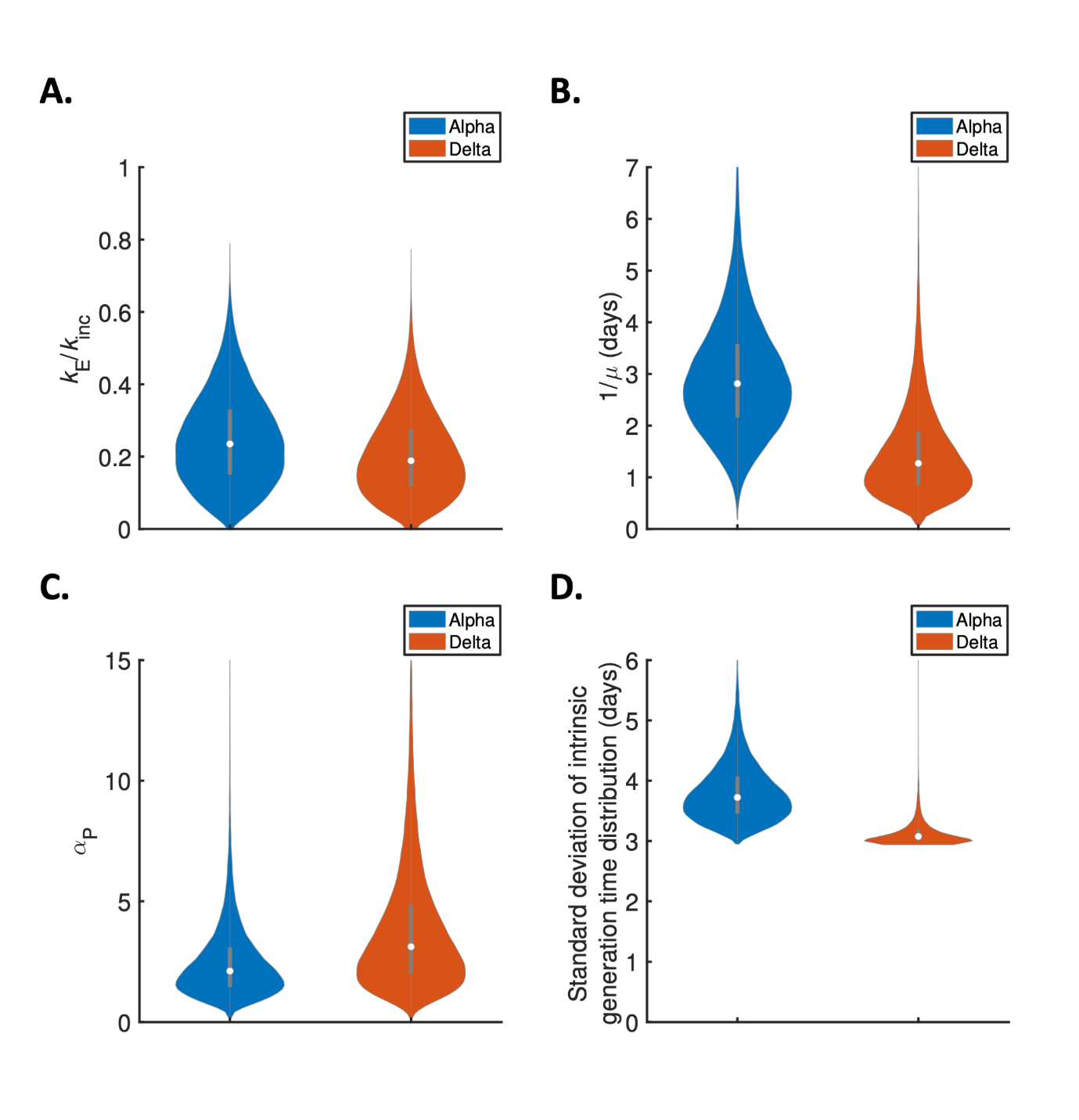

**Figure S1. Posterior distributions of fitted model parameters, and the standard deviation of realised generation times.** Violin plots indicate posterior estimates for the Alpha (blue) and Delta (red) variants of: A. The ratio of the mean durations of the latent (*E*) and incubation (combined *E* and *P*) periods, $k_{E}/k_{inc}$; B. The mean duration of the symptomatic infectious (*I*) period, $1/\mu$; C. The ratio of the transmission rates in the presymptomatic infectious (*P*) and symptomatic infectious (*I*) stages of infection, $\alpha_{P}$; D. The standard deviation of the intrinsic generation time distribution. Posterior means and 95% credible intervals for these quantities are given in Table S2 and Table S3.

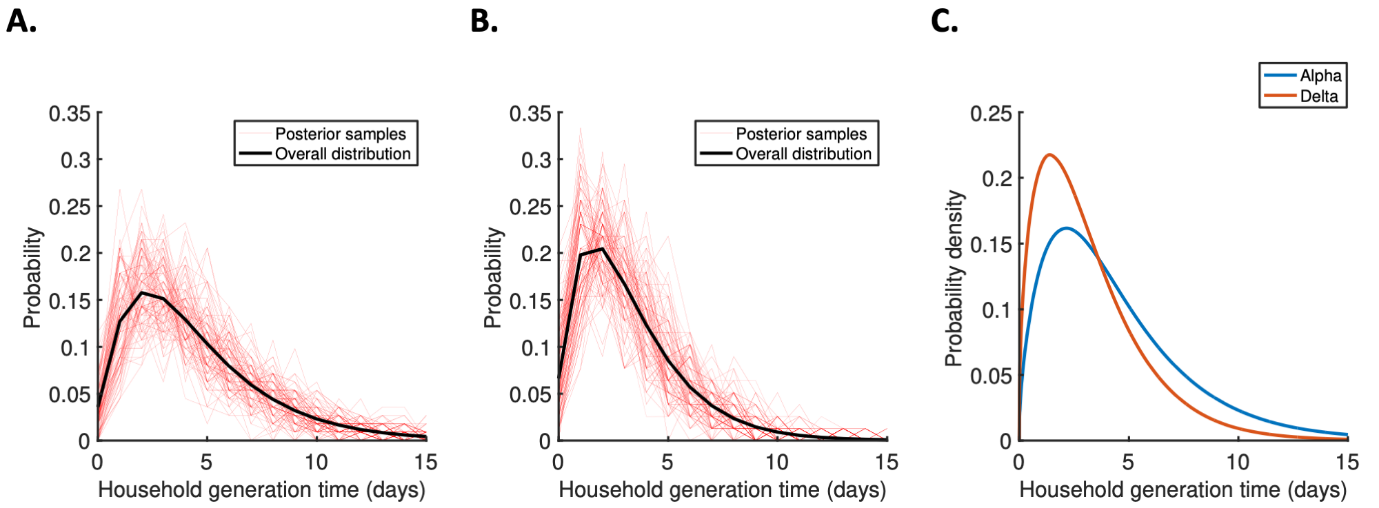

**Figure S2. Distributions of household generation times.** A-B. The discretised household generation time distribution (i.e., the number of days between the dates of individuals becoming infected and transmitting the virus) for the Alpha (panel A) and Delta (panel B) variants. In both panels, the red lines show the predicted distribution from 100 randomly selected steps of the MCMC procedure that we used to fit the mathematical model of household transmission to the UK household data, while the black line shows the overall distribution when combining the output of all MCMC steps (after burn-in and thinning). C. Comparison of the overall (continuous-time) household generation time distribution for the Alpha (blue) and Delta (red) variants.

### Supplementary Tables

| **Parameter** | **Interpretation** | **Value** | **Justification** |
| --- | --- | --- | --- |
| $\alpha_{A}$ | Relative infectiousness of entirely asymptomatic hosts | 0·35 | Taken from^1^ |
| $\eta_{j}$ | Relative susceptibility of individual $j$ (compared to an unvaccinated individual) | Dependent on the vaccination status of individual $j$:  1 (unvaccinated or within 20 days of first vaccine dose)  0·37 (Alpha variant; one dose of Oxford-AstraZeneca vaccine)  0·21 (Alpha variant; two doses of Oxford-AstraZeneca vaccine)  0·41 (Alpha variant; one dose of Pfizer-BioNTech vaccine)  0·22 (Alpha variant; two doses of Pfizer-BioNTech vaccine)  0·54 (Delta variant; one dose of Oxford-AstraZeneca vaccine)  0·33 (Delta variant; two doses of Oxford-AstraZeneca vaccine)  0·43 (Delta variant; one dose of Pfizer-BioNTech vaccine)  0·20 (Delta variant; two doses of Pfizer-BioNTech vaccine) | Consistent with previous estimates of vaccine efficacy against infection^2^ |
| $k_{inc}$ | Shape parameter of gamma incubation period distribution | 3·5 | Consistent with mean (5·8 days) and standard deviation (3·1 days) of previous estimates of incubation period distribution^3^ |
| $1/\gamma$ | Mean incubation period | 5·8 days | Consistent with previous estimates of incubation period distribution^3^ |
| $k_{I}$ | Shape parameter of (gamma) symptomatic infectious period distribution | 1 | Assumed |

**Table S1. Values of parameters that were not estimated from the household data**. All parameters listed were assumed to take the same values for the Alpha and Delta variants except where explicitly stated otherwise. No vaccine protection against infection was assumed for individuals who received their first vaccine dose fewer than 21 days before the first member of their household developed symptoms or returned a positive test, while increased vaccine protection from the second dose was assumed to take effect immediately (this was because no estimates for vaccine protection within three weeks of the first dose are given in^2^, whereas the estimates for protection within two weeks of the second dose from that study^2^ were similar to the estimates for protection after at least two weeks that we used here). Two individuals in our analyses who received the Moderna vaccine were assumed to have the same relative susceptibility as those who received the Pfizer-BioNTech vaccine. Two individuals who received an unspecified vaccine were assumed to have the same relative susceptibility as those who received the Oxford-AstraZenica vaccine.

| **Parameter** | **Interpretation** | **Prior** | **Posterior mean (95% CrI)** |
| --- | --- | --- | --- |
| $k_{E}/k_{inc}$ | Ratio of mean durations of the latent (*E*) and incubation (*E*+*P*) periods | Beta(2·1,2·1)  [prior median 0·5, 95% CrI 0·1-0·9] | Alpha: 0·25 (0·04-0·52)  Delta: 0·20 (0·03-0·46) |
| $1/\mu$ | Mean symptomatic infectious (*I*) period | Lognormal(1·6,0·8)  [prior median 5·0 days, 95% CI 1·0-23·8 days] | Alpha: 2·9 days (1·1-5·5 days)  Delta: 1·5 days (0·4-3·8 days) |
| $\alpha_{P}$ | Ratio of transmission rates in the presymptomatic infectious (*P*) and symptomatic infectious (*I*) stages | Lognormal(0,0·8)  [prior median 1·0, 95% CrI 0·2-4·8] | Alpha: 2·5 (0·7-6·7)  Delta: 3·9 (0·9-11·3) |
| $\beta_{0}$ | Overall transmissibility parameter | Lognormal(0·7,0·8)  [prior median 2·0, 95% CrI 0·4-9·7] | Alpha: 2·1 (1·7-2·5)  Delta: 4·4 (3·3-5·6) |

**Table S2.** **Values of fitted model parameters.** Descriptions of fitted model parameters, the prior distributions used (all prior distributions were taken to be independent, both between different parameters and between variants), and the posterior means and 95% credible intervals obtained for the Alpha and Delta variants.

| **Quantity** | **Posterior mean (95% CrI) for Alpha variant** | **Posterior mean (95% CrI) for Delta variant** |
| --- | --- | --- |
| Mean intrinsic generation time | 5·5 days (4·6-6·4 days) | 4·6 days (4·0-5·4 days) |
| Standard deviation of intrinsic generation time distribution | 3·8 days (3·1-5·1 days) | 3·1 days (3·0-3·7 days) |
| Mean household generation time | 4·5 days (3·7-5·4 days) | 3·2 days (2·4-4·2 days) |
| Standard deviation of household generation times | 3·3 days (2·6-4·2 days) | 2·4 days (1·8-3·1 days) |

**Table S3.** **Mean and standard deviation of the intrinsic and household generation time distributions.** Posterior means and 95% credible intervals for the mean and standard deviation of the intrinsic and household generation time distributions for the Alpha and Delta variants.
